# Protocol for the development of reporting guidance for interest-holder engagement in practice guidelines: the RIGHT-MuSE checklist

**DOI:** 10.1101/2025.02.13.25322255

**Authors:** Xuan Yu, Janne Estill, Elie Akl, Joanne Khabsa, Jennifer Petkovic, Lili Zeidan, Zhaoxiang Bian, Yaolong Chen

## Abstract

**Introduction:** Guideline developers have increasingly engaged various groups of interest-holders in different stages of guideline development. However, reporting on interest-holder engagement in practice guidelines often lacks transparency. Therefore, the RIGHT (Reporting Items of Practice Guidelines in Healthcare) working group, in collaboration with the MuSE Consortium, plans to develop the RIGHT-MuSE extension.

**Objectives:** This protocol aims to outline the steps for the detailed development of the RIGHT-MuSE checklist, as well as strategies for its effective development, dissemination, and application.

**Methods:** We will follow the methods recommended by the EQUATOR (Enhancing the QUAlity and Transparency Of health Research) Network, and build on the experience from the development of the original RIGHT statement and its extensions. The development process of RIGHT-MuSE will consist of twelve specific steps: 1. Identifying the need for the checklist; 2. Obtaining funding; 3. Drafting a protocol and registering the project; 4. Establishing the working groups; 5. Reviewing background work; 6. Generating an initial list of items; 7. Conducting consensus surveys; 8. Holding consensus meetings; 9. Drafting the final RIGHT-MuSE checklist; 10. Conducting a pilot test of the checklist; 11. Developing an explanatory document; and 12. Disseminating the checklist.

**Discussion:** The RIGHT-MuSE checklist will provide guideline developers with guidance for the systematic, scientific and transparent reporting of interest-holder engagement in practice guidelines. Additionally, developers and implementers of the RIGHT-MuSE checklist will use this protocol as a reference to ensure that the checklist they develop and implement adheres to the highest standards of transparency and quality. By promoting active and meaningful engagement of interest-holders, the RIGHT-MuSE checklist aims to foster inclusive and people centered processes in healthcare.

## Introduction

In recent years, guideline developers have increasingly engaged various interest-holder (commonly also known as ‘stakeholder’[1]) groups—particularly patients and the public—to participate in all stages of guideline development[2][3]. Organizations involved in guideline development and dissemination, such as the World Health Organization, the National Institute for Health and Care Excellence, and the Guidelines International Network, recommend the consideration of interest-holder engagement throughout the guideline development process[4][5][6]. Such engagement not only enhances the consideration of contextual factors in the guideline development process but also promotes ownership and uptake among the end users.

The MuSE (formerly Multi-Stakeholder Engagement) Consortium is an international network of over 160 individuals interested in engagement in research and guidelines[7]. Currently, the MuSE Consortium has begun to adopt the term “interest-holders” to describe groups with legitimate interests in the health issue under consideration[1]. Enhanced engagement of interest-holders can result in better informed decision-making about the selection, conduct, and use of research[8]. Therefore, this project will use the term “interest-holder engagement”.

Although guideline developers are increasingly considering engaging representatives of all interest-holder groups in the development process, reporting on the engagement of interest-holders often lacks transparency[2]. Questions such as which interest-holders were included, how they were recruited, how they were engaged in the process, and whether equity considerations were addressed, are often not described in detail. The RIGHT (Reporting Items for practice Guidelines in HealThcare) checklist[9] is a comprehensive reporting tool for practice guidelines in healthcare, and it includes two items related to the involvement of interest-holders (items 9a and 9b). However, these two items only cover a description of how all contributors to the development of the guideline were selected, their roles and responsibilities, as well as a list of all individuals involved in developing the guideline.

To address these gaps, we plan to develop a standardized reporting tool for interest-holder engagement in practice guidelines (RIGHT-MuSE extension). It aims to help guideline developers to systematically, scientifically, and transparently report how all interest-holders were engaged throughout the guideline development process. This protocol outlines the steps for developing the RIGHT-MuSE checklist in detail, including our plan for inviting relevant interest-holders to participate in this process, and strategies for the checklist’s effective development, dissemination, and application.

## Methods

We will develop the RIGHT-MuSE reporting guidelines following the methods recommended by the EQUATOR (Enhancing the QUAlity and Transparency Of health Research) Network[10] as well as the experience from the development of the original RIGHT statement and its extensions[9][11][12][13][14][15]. The RIGHT-MuSE project has been reviewed and approved by the Ethics Review Committee of Lanzhou University Institute of Health Data Science, China (HDS-202412-01). The development process of RIGHT-MuSE will consist of five stages encompassing twelve specific steps. Table 1 shows the development process and the anticipated timeline. In the following sections, we will present each step in detail.

**Table 1.**
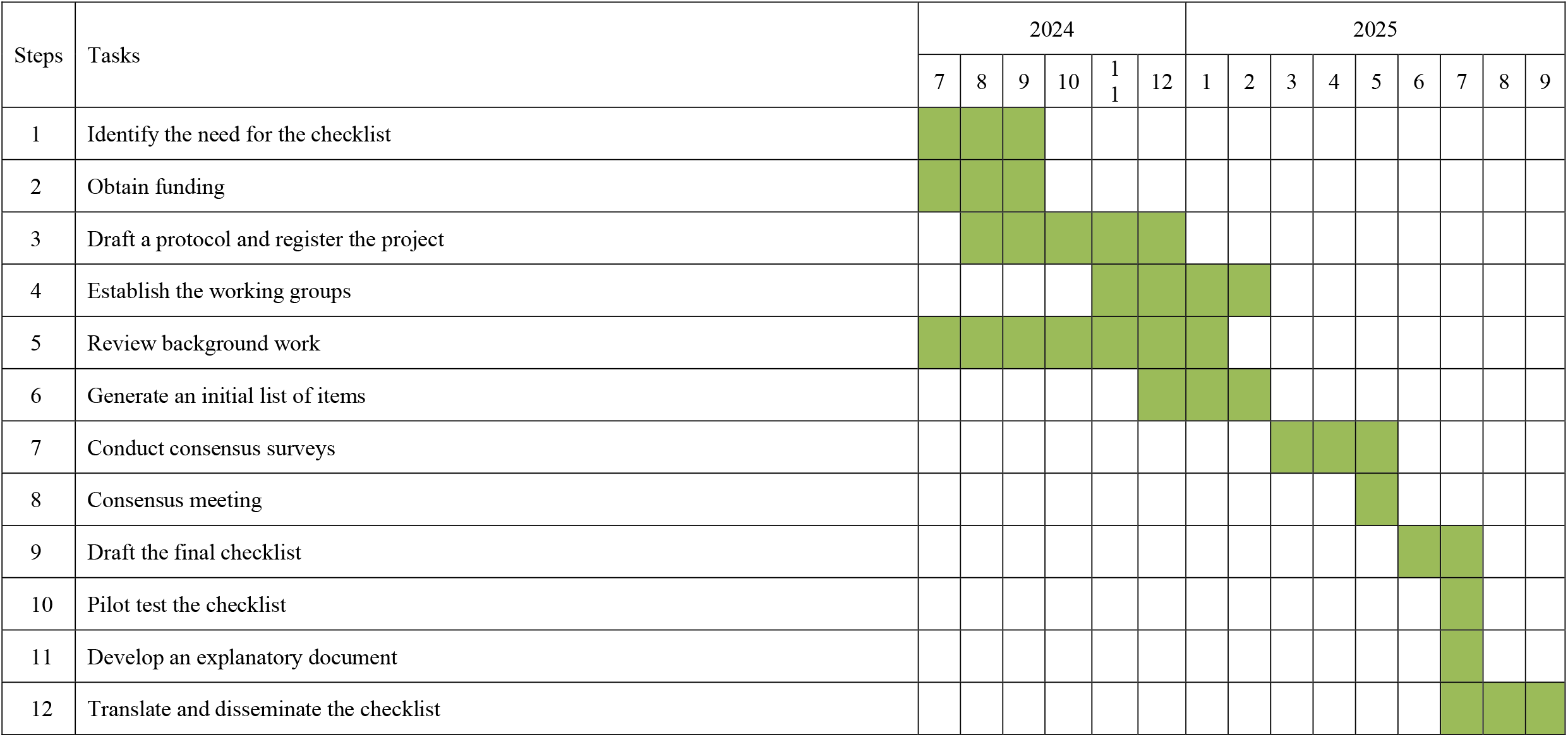
Process and timeline of the development of the RIGHT-MuSE reporting guideline.

### 1. Identify the need for the checklist

After a thorough review of published literature and reports, we did not find any published guideline on how to scientifically and transparently report the engagement of interest-holders in the development of practice guidelines. Further consultations and discussions with experts have led to a consensus on the importance of developing such a checklist. Therefore, we have decided to develop an extension of the RIGHT reporting guideline for interest-holder engagement—the RIGHT-MuSE extension.

### 2. Obtain funding

This project is funded by the Vincent and Lily Woo Foundation and CAMS Innovation Fund for Medical Sciences — Research Unit of Evidence-based Evaluation and Guidelines (2021RU017). The funders have no role in the study design, data collection and analysis, manuscript writing, or publication process.

### 3. Draft a protocol and register the project

The protocol was drafted following the methodology recommended by the EQUATOR Network[10] and the methods used in the development of the original RIGHT statement and its other extensions[9][11][12][13][14][15]. The protocol has been registered with the EQUATOR Network (https://www.equator-network.org/library/reporting-guidelines-under-development/reporting-guidelines-under-development-for-other-study-designs/#RIGHTMuSE).

### 4. Establish the working groups

The RIGHT-MuSE working group will comprise experts from various disciplines (e.g., health sciences, social sciences, and journal editors). We will aim to ensure broad geographical diversity and representation of different research backgrounds, including evidence-based medicine, reporting guideline development, equity, and evidence-based social sciences. We will also invite members from the working groups of the RIGHT statement and its previous extensions to participate. The RIGHT-MuSE working group will be divided into two subgroups: the Coordination team and the Consensus panel.

All members involved in RIGHT-MuSE will be required to declare their interests.

#### Coordination team

The coordination team will consist of 7 members (XY, JE, EA, JK, JP, LZ, and YC) with experience in project management, development of reviews and reporting guidelines, and conducting qualitative and quantitative research. The specific roles of the coordination team are to: (1) manage the project timeline and process; (2) draft and submit the final protocol for registration with the EQUATOR Network; (3) establish the consensus panel; (4) obtain and manage the conflicts of interest for all group members; (5) conduct the necessary literature reviews; (6) generate the initial list of items; (7) organize consensus surveys and analyze the results; (8) organize and facilitate the consensus meetings; (9) draft the final checklist and explanatory document; (10) test the checklist; and (11) prepare the checklist for publication.

#### Consensus panel

The consensus panel will consist of around 20 globally recruited representatives of different interest-holder groups. We will align the recruitment with the classification of interest-holders in guideline development proposed by Petkovic *et al*. which contains ten categories: patients (including patient caregivers and patient advocates/organizations), payers of health research, payers/purchasers of health services, editors of peer-reviewed journals, policymakers, principal investigators and members of research teams, product makers, program managers, providers in healthcare, and members of the public[2]. Among them, at least 10 experts will be selected from the group of principal investigators and research team members who have experience in practice guideline development, reporting guideline development, and research related to interest-holder engagement, equity, human rights, or gender issues. These experts will also include representatives of the Guidelines International Network (GIN), the RIGHT working group, the Appraisal of Guidelines for Research and Evaluation (AGREE) collaborative organization, the Grading of Recommendations Assessment, Development, and Evaluation (GRADE) working group, the MuSE Consortium, and other relevant organizations and guideline interest-holders. The key tasks of the consensus panel include: (1) contributing to the selection and identification of items to be included in the final RIGHT-MuSE checklist through participation in consensus surveys and consensus meetings; and (2) reviewing, revising, and approving the final RIGHT-MuSE checklist and explanatory document.

### 5. Review background work

To collect the initial list of items for RIGHT-MuSE, we will review the findings of a methodological survey of interest-holder engagement approaches outlined in methodological guidance documents of guideline-producing organizations[16], the findings of a scoping review of the guidance for engagement in health guideline development[17], and the GIN-McMaster guideline development checklist extension for engagement[18]. We will also refer to relevant articles published by the MuSE Consortium regarding interest-holder engagement in practice guidelines[2][19][20][21]. These studies will help to clarify the currently used processes for identifying, recruiting, and selecting interest-holders to engage in practice guidelines and collect information on the methodology used in reporting interest-holder engagement in practice guidelines.

### 6. Generate an initial list of items

Two members of the coordination team (XY and LZ) will independently collect a pool of potential items based on the work described in the previous step and will agree on items to be presented to the coordination team. Subsequently, all members of the coordination team will hold several rounds of meetings to determine whether the initial list of items is comprehensive enough, as well as the importance, applicability, accuracy, and clarity of each item. The coordination team will record the discussions and document the reasons for any modifications, additions, or deletions to the items, until a version of the checklist suitable for the consensus process is reached.

### 7. Conduct surveys

We will conduct one or more rounds of online surveys (using methods similar to the Delphi process) as needed to reach consensus among the consensus panel regarding the items to be included in the RIGHT-MuSE checklist. During the first round of the survey, we will provide the project description to the panelists, collect informed consent forms and demographic information of the panelists, and conduct the survey of the initial items. In the survey, we will present each item and its explanation. Panelists will provide their perspectives on whether each item should be kept without changes, modified, or excluded. Additionally, the panelists will be asked to provide any comments on existing items or suggest new items to be included. We will use a 66% agreement threshold as a guide to decide on keeping an item without changes or excluding it[22][23]. If all items received consensus in the first round of the survey and there are only a few comments and suggestions, we will end the consensus survey and proceed directly to the consensus meeting.

In the second round of the survey, the coordination team will provide the results and feedback from the first round of the survey. Panelists will be invited again to provide their views on the updated items. Items for which no consensus is reached will be discussed at the next step (consensus meeting).

### 8. Consensus meeting

After completing the surveys, we will hold a consensus meeting to present and discuss the survey process and results. We will invite all members of the coordination team and consensus panel to participate in this meeting. The consensus meeting is planned to be conducted online; if it is not possible to find a time suitable for everyone, we will hold 2-3 meetings at different times to accommodate as many participants as possible. The meeting will be chaired by a representative from the collaboration team, and will record the meeting proceedings. After the meeting, the collaboration team will compile all suggestions and comments mentioned during the discussions and provide feedback via email within five working days.

### 9. Draft the final RIGHT-MuSE checklist

After completing the consensus meetings, a final version of the RIGHT-MuSE checklist will be drafted by the collaboration team. All members of the consensus panel will be invited to review the final version of the checklist.

### 10. Pilot test of the checklist

The items will be pilot-tested using both human evaluators and large language models (LLMs), such as ChatGPT-4o. We will compare the consistencies between human evaluators and LLMs, different humans, and different input queries to LLMs. First, we will invite two guideline evaluators (who were not part of the RIGHT-MuSE working group) to work in pairs and independently evaluate the compliance of ten practice guidelines to the RIGHT-MuSE checklist. A Kappa value greater than 0.7 will indicate good consistency in the understanding of the items between the evaluators, thus affirming the reliability of the evaluation results. We will use these results as a reference standard for the subsequent steps. Then, the collaboration team will train ChatGPT-4o on all items and evaluate the same ten practice guidelines. If the accuracy rate of the LLM remains below 70% after three to five rounds of adjustment to the instructions, we will consider that there are issues with the description of items and revise them to ensure there is no confusion. Conversely, if the accuracy rate is 70% or higher, the content of the items will be deemed clear. When all items achieve an accuracy rate of at least 70%, the RIGHT-MuSE checklist will be considered to have good consistency. Finally, we will also use different LLMs to evaluate the items. Each model will be tested using a standardized set of queries designed to elicit responses regarding the content of the items. The outputs from each LLM will be collected and analyzed for consistency. If the Intraclass Correlation Coefficient value exceeds 0.7, it is considered to indicate a good level of consistency among the different models. When all pilot tests meet the expected results, the final version of the RIGHT-MuSE checklist will be considered ready for submission for publication.

### 11. Develop an explanatory document

In addition to the RIGHT-MuSE checklist, an explanatory document for the users of the checklist will be developed. The document will include a detailed explanation and a statement of the rationale for each checklist item, and examples of good reporting.

### 12. Disseminate the checklist

We will disseminate the RIGHT-MuSE checklist in various ways to ensure widespread awareness and adherence during practice guideline development. We plan not only to publish the RIGHT-MuSE statement in a peer-reviewed journal but also to make it available on the EQUATOR Network and its branches (such as the Chinese EQUATOR Centre), the RIGHT website, the MuSE website, and through other relevant guideline-related organizations. Additionally, we will conduct workshops and present the checklist at relevant conferences to further promote its use. Furthermore, we encourage our contributors, partners, and other researchers to translate this reporting guideline into different languages to facilitate its adoption in various countries and regions.

## Discussion

The primary outcome of the RIGHT-MuSE project will be the establishment of a standardized checklist for interest-holder engagement in guideline development, and an accompanying explanatory document. The collaboration team of the RIGHT-MuSE working group will compile and organize all materials collected throughout the checklist development process. Once the checklist is finalized and published, we will actively promote and implement it to foster the development of more trustworthy practice guidelines.

As an extension of the RIGHT reporting guideline, RIGHT-MuSE will build upon existing items to focus specifically on reporting interest-holder engagement throughout the guideline development process. Currently, researchers are actively developing guidance for interest-holder engagement in the creation and implementation of practice guidelines and analyzing facilitators and barriers to interest-holder engagement, which emphasizes the importance of developing RIGHT-MuSE[2][19][20]. The collaboration between the RIGHT and MuSE working groups, application of the rigorous methodology recommended by the EQUATOR Network, and the particular focus on equity in the development process will guarantee the high quality of the RIGHT-MuSE reporting guideline.

We hope that the publication of this protocol will help us recruit a multidisciplinary team of different interest-holders to join the Consensus panel for this project, provide diverse perspectives and opinions, and offer valuable recommendations and insights for the future dissemination and updating of RIGHT-MuSE.

## Author contributions

*Conceptualization:* Elie Akl, Zhaoxiang Bian, and Yaolong Chen. *Methodology:* Xuan Yu, Janne Estill, Elie Akl, Joanne Khabsa, Zhaoxiang Bian, Yaolong Chen, Jennifer Petkovic, Lili Zeidan. *Writing—original draft:* Xuan Yu, Zhaoxiang Bian, Janne Estill. *Writing—review & editing:* Xuan Yu, Janne Estill, Elie Akl, Joanne Khabsa, Zhaoxiang Bian, Yaolong Chen, Jennifer Petkovic, Lili Zeidan.

## Data availability statement

All data from this protocol are available upon reasonable request. Please contact the corresponding authors for details.

## Ethics approval statement

The RIGHT-MuSE project has been reviewed and approved by the Ethics Review Committee of Lanzhou University Institute of Health Data Science, China (HDS-202412-01).

